# Would India Really Touch the Peak of SARS COVID 19 Cases or Deaths in Near Future?

**DOI:** 10.1101/2020.05.21.20109728

**Authors:** Pramod K. Gupta

## Abstract

**Background:** The Government, Health System and even an individual citizen of India is alarmed expecting the height of pandemic of SARS-COVID-19 in near future. Many experts worldwide predict it to happen in India between end of May and end of July.

**Objectives:** The aim of this research was to find an answer that whether India would come across the looming conditions of SARS-COVID-19 in coming days given the prevailing circumstances so far.

**Methods:** The proposed approach used fundamental concept of Statistics by fixing the standard reference to the number of daily new tests conducted by a country. We thus computed the percentage of daily new cases and daily new deaths, in using such references. The trends were studied using simple line chart. The theory of three sigma was also used to build the upper bound for daily new cases and deaths, specifically for India to see the extreme conditions.

**Results:** The analysis was done using data from January to till May 18, 2020 for India, Italy, USA and UK. The trend of India was almost fix between ~2% to ~6% till May 18, 2020. On contrary, Italy, USA and UK were touched the Peak on March 29, 2020 (24.38%), April 26, 2020 (23.51%) and April 24, 2020 (24.91%), respectively and declining since then. Similar trends were also noted in daily new deaths, except Italy.

**Conclusions:** The proposed new concept for fixing universal reference provides a consistent and coherent results. It is thus clear from observed data so far that India is not going to encounter the frightened conditions or peak, like, Italy, USA, and UK for pandemic SARS-COVID-19, given the existing conditions, excluding the current migration.

## Introduction

The ongoing pandemic of SARS-COVID-19 is affecting the world worst even after a couple of months. Many questions and doubts related to SARS-COVID-19 are not running only in general population, Government but also among the experts. India is 2nd largest in population in the world and located in the close proximity of China. However, the nature of the SARS-COVID-19 pandemic is not like as being seen in the countries like Italy, UK, USA, etc., till today, but the expectation of the worst in coming days is looming large. Many experts opine that India will see the peak of the SARS-COVID-19 by the end of May/June/July.

As far our search of literature concern, no peer review study so far seen that directly address the question raises here. A few, precisely one publication on good preprint server was found dealing with prediction of cases in India, which result is far from practicality. These non-peer reviewed publications are mostly based on SIR model, moving average modal, some trend analysis for number of cases India will touch/double on specific date, etc., whereas notes and commentary from the experts on print and electronic media are based on same time dependent trend theory or simply an opinion lacks scientific contents to referred herein. These trend analyses depend on choice of time, like moving average. However, Time dependent concept governs by the wisdom of researcher to choose the time frame and thus lack the coherency. If the processes have different onset time then it also leads difficulty in fair comparison and utilization of data.

We aimed to investigate the question whether India would have peak in coming time and compared the situations with other countries by proposing a new approach that does not depend on time frame.

## Methods

The proposed study is theoretical in nature and the proposed new approach were fit with the available SARS-COVID-19 data available in public domain. It was therefore, standard set-up of study design, sampling approach, inclusion/exclusion criteria, etc., not needed to describe. The new simple approach based on the logic of fundamental concept of Statistics for estimating/predicting the change with respect to a specific reference point and the theory of independent identical random sampling as well. To understand the true burden of pandemic, daily new SARS-COVID-19 tests were performed by every country, including India. We considered that the number of daily new tests done by any country as a study sample, which represent the population under risk for SARS-COVID-19. We thus computed the percentage of daily new positive cases keeping number of daily new test done as a reference. We used the same approach to obtain the percentage of daily new deaths due to SARS-COVID-19 changing the reference point to daily new positive cases instead of daily new tests.

We thus used these computed percentages to plot a simple line chart for selected countries to see the trend. The proposed approached was applied using data of daily new test done, daily new positive cases and daily new deaths reported by India, Italy, USA and UK. The selection of countries was done purposefully. Italy was considered because the effect of the SARS-COVID-19 appeared climbing down, USA & UK were considered because the effect of the SARS-COVID-19 still recorded on high. These countries were seen in contrast to the India where the worst of the SARS-COVID-19 expected to hit in near future. The summary statistics, Mean and SD by omitting 02-data points of the beginning dates were also computed further for percentage daily new cases and deaths, exclusively for India only. The theory of 3-sigma was thus used to obtain the upper bound for percentage daily new cases and deaths. The said upper bound gave statistical meaningful condition for understanding the extreme circumstances.

## Data

We used the data available on the website “Our World in Data”^1^ downloaded on the May 18, 2020. Many raw as well as analyzed data files were available, which were being routinely updated. We downloaded CSV files of Raw data related to number of daily new test, number of daily new cases and number of daily new deaths recorded for all countries on May 18. 2020. The data were synthesized and prepared for further analysis without any alteration. Data for different countries started from different dates and were also not available for some of the dates.

## Result and Discussion

The results depicted in Figure-01 to Figure-04, were respectively comprised the trend for India, Italy, USA and UK. The data was consistently available from the month of January for Italy but it was not consistent for India, USA and UK till the end of February. Hence, the trend depicted in figures was not quite consistent during these months for India, USA and UK. It is also to mentioned that the data was primarily missing during these said periods for all countries but Italy. It was thus important to point out that the trend lines related to daily new cases or deaths, whenever it touched the x-axis implied unavailability of data on that particular date.

**Figure-1:**
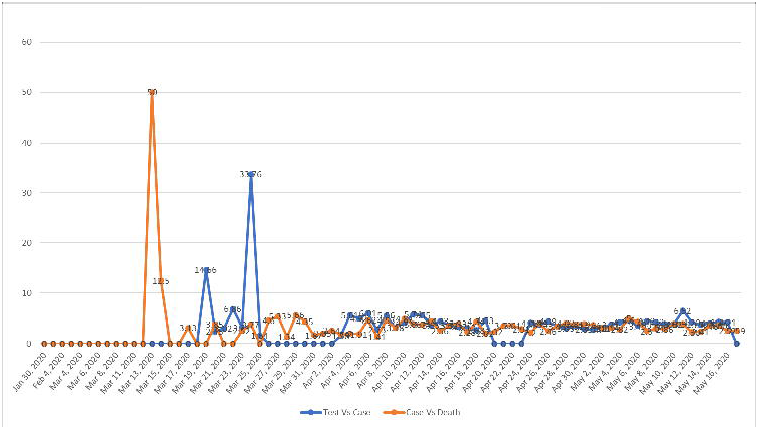
Trend (%) of SARS-COVID-19 Cases and Deaths For India

**Figure-2:**
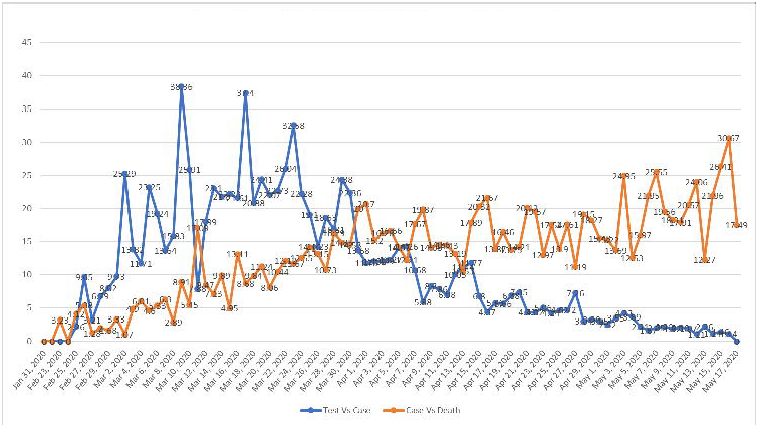
Trend (%) of SARS-COVID-19 Cases and Deaths For Italy

**Figure-4:**
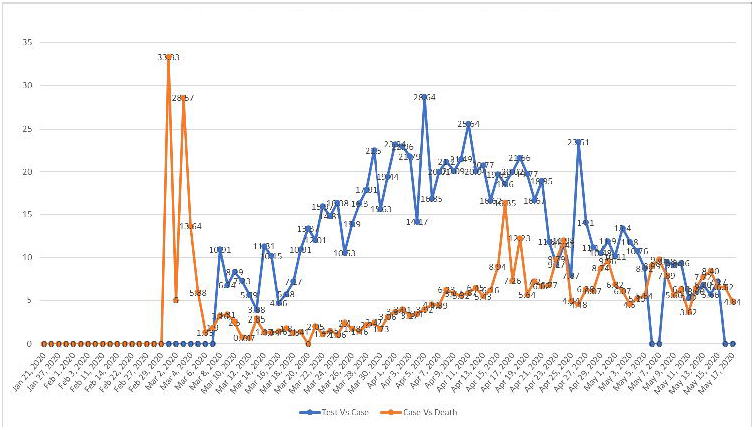
Trend (%) of SARS-COVID-19 Cases and Deaths For U K

However, it was very clear from the Figure-01 to 04 that the percentage of daily new cases in reference to daily new tests of SARS-COVID-19 showed noticeable changes over time (Figure-02 to 04) in case of Italy, UK and USA. It can also be noticed from the Figure-02 that after March 29, 2020 (24.38%) the new cases were climbing down till May 16, 2020 (1.14%) for Italy. In case of USA, and UK, the similar conditions can be seen on April 26, 2020 (23.51%) and April 24, 2020 (24.91%), respectively, which were going down till May 16, 2020 within range of ~10% to ~5%. and May 16, 2020 (~5%), respectively. However, the trends for USA and UK were not as explicit as for Italy. The trends for Italy, USA and UK shown many spikes during different intervals and percentage went up to threating level. It was clear indication of the calamity of SARS-COVID-19 pandemic. The tale of daily new deaths in these countries were, shown in the same Figure-2 to 4, also giving chillness to spine. Italy though showed clear downward trend, recording daily new cases down to 1.14% but the trend of daily new deaths varied over ~10% to over 30%.

On contrary, the trend of India shown in Figure-1 was almost constant over time, precisely varied between ~2 to ~6 percent. The trend of new cases stands in certain range as mentioned before, considering total daily new testing done by India over 40-thousands since May 24, 2020. Over 40-thousand daily new testing appeared to be justified given the simple logic of sampling that was beyond the scope this work. However, these findings were empirically proved given the considered case of Italy, USA and UK, where number of daily new positive cases still turned higher, whether total number of daily new test were larger or smaller (not too small number of tests, like less than 15-20 thousand). It happened because the positive cases were present within the population under risk. The situation was not being encountered in India. Hence, fear of spike would be seen in India appeared to be false given the data recorded so far. The further analysis of percentage data was done for India and summary statistics were obtained. The average (Mean) and dispersion (SD) were obtained for daily new cases (mean: 3.87; sd: 1.15) and deaths (mean: 3.33; sd: 1.05). Three sigma rules for upper bound turned out 7.33% for daily new cases and 6.48% for daily new deaths. These percentages would increase or decrease given the number of new test decrease or increase respectively, as it was noted very clearly on May 11, 2020 when total daily new test was (64651) low than the percentage of new cases went up to 6.52%, whereas this percentage went down to 3.72% for daily new test done 94671. Henceforth, it was quite obvious that the so-called peak of SARS-COVID-19 would be somewhere around ~7.33%. It can be considered coherent as the SARS-COVID-19 pandemic stepped into 5^th^ month, almost equal duration of Italy and other countries, where its effects were going down in data. The trend of percentage daily new deaths for India was same as per the trend of daily new cases as revealed by Figure-1, which were also similar to USA and UK. However, the magnitude of percentage for USA and UK was recorded much higher than India

## Conclusions

The proposed method is based on new concept that uses fundamental Inferential statistical theory and theory of sampling as well. The propose new standard reference point, which is number of daily new tests done by any country with respect to daily number of new positive cases and deaths. The results are obtained for Italy, USA and UK to address the problem in hand with respect to India turn to be consistent, coherent and matches the real conditions. Hence, the trend for India based on used data and given prevailing clearly shows no spike. The result is persistent in trend and explains it well within the domain of statistics. However, it is obvious that the number of daily new cases as well deaths keep increasing with the shown percentage. The daily new positive cases may rise up to ~7.5% given the daily new testing range between 85-95 thousand. It is also pertinent to mention that the findings for India are limited for existing data and ongoing prevailing conditions, which excludes the current migration status of people from one state of India to other. The findings should not be taken for granted for giving relaxation in the near future and thus maligned the strict measures and sacrifices done so far.

It is also very important to point out Italy for its trend of daily new deaths. It is highly alarming and the country should take strict measures to control the current situation. If the countries USA and UK withhold and improve upon the measures that are being taken so far, for curving the SARS-COVID-19, thus the ongoing trend of daily new cases as well as daily new deaths for UK and USA may decline in the coming time too.

## Data Availability

All data is given in the manuscript

https://ourworldindata.org/grapher/daily-cases-covid-19

## Acknowledgment

The author is thankful to those who manages website of “Our World in Data” for their efforts and routinely maintaining the world wide data related to SARS-COVID-19.

## Contributors

PKG conceptualized, searched & retrieved data, performed analysis, wrote, revised the manuscript for worthy content.

## Financial support & Sponsorship

NO grant whatsoever is received for this study.

## Conflicts of Interest

None declared.

## Patient and Public Involvement

No patient and public involved.

## Ethics Approval

The research is completely theoretical and the data involved therein is obtained from public domain available without any restriction to download. Hence such approval is NOT required.

## Provenance and Peer Review

Not commissioned; externally peer reviewed.

## Data Availability Statement

The data used in this study is publicly available and which detailed is already given in this manuscript.

## Notes

### Competing Interest Statement

The authors have declared no competing interest.

## Reference

1. https://ourworldindata.org/grapher/daily-cases-covid-19

